# Predictors of Depression and Anxiety Symptoms in Brazil during COVID-19

**DOI:** 10.1101/2021.06.28.21259409

**Authors:** Stephen X. Zhang, Hao Huang, Jizhen Li, Mayra Antonelli-Ponti, Scheila Farias de Paiva, José Aparecido da Silva

**Author notes:** Correspondence; 9-27 Nexus10 Tower, 10 Pulteney St, Adelaide SA 5000, Australia.

## Abstract

The COVID-19 pandemic in Brazil is extremely severe, and Brazil has the third-highest number of cases in the world. The goal of the study is to identify the prevalence rates and several predictors of depression and anxiety in Brazil during the initial outbreak of COVID-19. We surveyed 482 adults in 23 Brazilian states online on 9–22 May 2020, and found 70.3% of the adults (N=339) had depressive symptoms and 67.2% (N=320) had anxiety symptoms. The results of multi-class logistic regression models revealed that females, younger adults and those with fewer children had a higher likelihood of depression and anxiety symptoms; adults who worked as employees were more likely to have anxiety symptoms than those who were self-employed or unemployed; adults who spent more time browsing COVID-19 information online were more likely to have depression and anxiety symptoms. Our results provide preliminary evidence and early warning for psychiatrists and healthcare organizations to better identify and focus on the more vulnerable sub-populations in Brazil during the ongoing COVID-19 pandemic.

## 1. Introduction

Since the initial outbreak of COVID-19 in late 2019, the COVID-19 epidemic has caused a devastating blow to the world, including the death of millions of people and the setback of socioeconomic functions of society and individual daily life [1]. It was reported that 2.6 billion people experienced emotional and economic shocks; this number even exceeds the number of people affected by the Second World War. Brazil has one of the highest rates of COVID-19 cases and deaths in the world by so far. Since the beginning of 2020, the pandemic has had a great impact on the normal life of people in Latin America [2-4], including its largest country Brazil [5,6]. The COVID-19 pandemic not only threatens people’s health but also impacts on the mental health of the public [7-10]. On the one hand, the uncertainty of the initial route of transmission and treatment has exacerbated people’s fear during the COVID-19 crisis [11]. On the other hand, the social distancing and confinement measures during the COVID-19 pandemic can lead to symptoms of anxiety and depression [12-17]. Although several studies have documented high prevalence rates of mental health symptoms in various parts of the world, especially in China [8,18-20], and early evidence of the mental impact of COVID-19 in Brazil was reported in a timely manner [21], there are few evidence-based studies containing critical predictors of mental issues in adults under a full-on COVID-19 outbreak in Brazil. As the devastating COVID-19 crisis continues in Brazil, it is crucial and urgent to investigate the risk factors for mental health issues in Brazilian adults.

In this study, we use three types of predictors: demographic factors, health factors and COVID-specific factors. The research on mental health in the COVID-19 pandemic has focused on demographic characteristics as predictors [22-24]. For example, gender [25-27], age [28], education [29], occupation [30] and number of children [31] have been the key predictors. Based on the existing research, we examined the key demographic factors such as gender, age, education, occupation and number of children as the predictors in this study. Good health and behaviors such as exercise [8] can improve individuals’ mental health. On the contrary, individuals with chronic health problems are more likely to encounter mental disorders [32]. Hence, we examined exercise and the presence of chronic diseases as predictors of mental health under the COVID-19 pandemic. Of COVID-19 specific factors, the emerging literature on COVID-19 mental health has uncovered that the occurrence of symptoms related to or similar to COVID-19 increases individuals’ psychological risk [33]; and information on COVID-19 on the internet is an important predictor of people’s fear or panic over COVID-19 [26]. Therefore, we include the symptoms of COVID-19 infection and the hours per day spent browsing COVID-19 information online as COVID-19-related predictors in this study.

This study attempts to identify the predictors of depression and anxiety symptoms of Brazilian people during the COVID-19 pandemic to provide preliminary evidence and early warning for psychiatrists and healthcare organizations to identify the more vulnerable sub-populations to enable more targeted and timely mental intervention.

## 2. Materials and Methods

### 2.1. Design

We launched an online survey to study the mental health of adults in Brazil during the COVID-19 pandemic on 9–22 May 2020. The survey was conducted through a link to Google Forms to preserve social distancing protocols and to reach people across Brazil’s large and diverse regions. We used the non-probabilistic sampling technique of quota sampling to approximate a representative sample of Brazilian adults. Quota sampling is one of the most popular sampling methods and a viable method [34] to conduct online surveys across all regions of Brazil without access to a probabilistic panel. The use of quota sampling by age, gender and social class was effective and viable in our case to obtain a sample that represents the population in Brazil. The study sampled adults aged 18 years or older by unclustered systematic random samples from all 23 states in Brazil. The survey, in Brazilian Portuguese, contained a cover page, which explained the purpose of the study and all the participants consented before starting the survey. From the 857 adults who participated in the study, we received a total of 482 valid responses for a response rate of 56.2%. Ethical approval for this research was received from the Ethics Review Board (CAAE: 31703720.9.1001.0008) at University of São Paulo.

### 2.2. Variables and instruments

The survey collected socio-demographic information of individual adults, including their gender, age, education level, employment status, work and family status. The survey also collected basic health conditions such as chronic health issues [8] and lifestyle behaviors such as daily exercise time [33]. The descriptions of these variables are in Table 1.

**Table 1.**
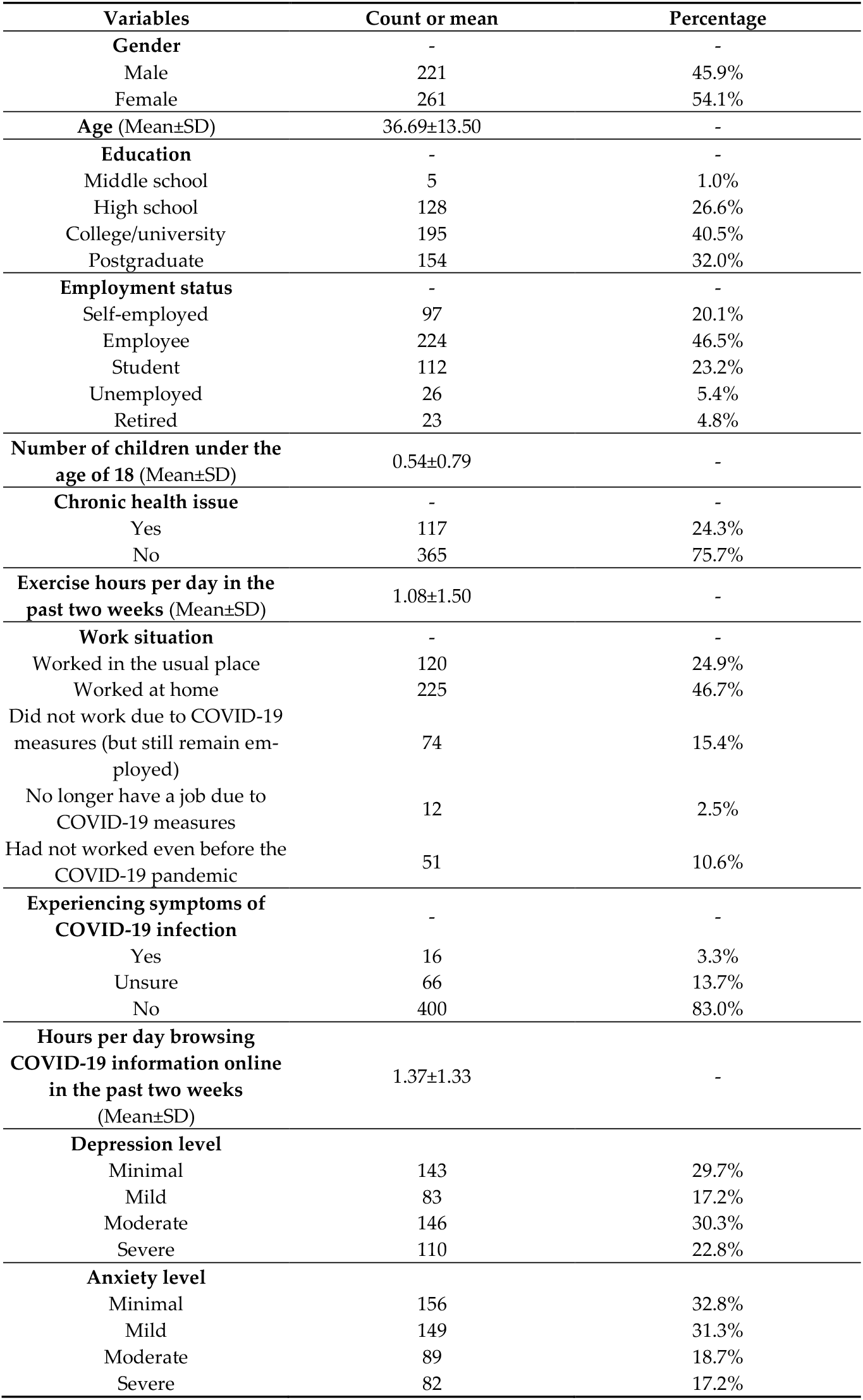
Descriptions of the participants (N=482).

The outcome variables are depression [35] and anxiety [36]. The Patient Health Questionnaire-9 (PHQ-9) is one of the most established depression scales, which captures the frequency and severity of depression related symptoms in the past two weeks, with a total of 9 items. In this study, the internal consistency coefficient of PHQ-9 is 0.902. A sample item is: in the past two weeks, how many days did you have a lack of appetite or did you overeat (0 = no day 1 = less than a week 2 = a week 3 = almost every day)?

The Generalized Anxiety Disorder scale (GAD-7) is a simple and effective way to evaluate generalized anxiety disorder (GAD), a mental disorder with long-term persistent anxiety and excessive anxiety as the core symptoms. In this study, GAD-7 has 7 items, and the internal consistency coefficient is 0.937. A sample item is: in the past 2 weeks, how often did you feel nervous, anxious or very tense (0 = rarely 1 = some days 2 = more than half the days 3 = almost every day)?

### 2.3. Statistical strategy

All the data processing was completed in SPSS 23.0, and a two-tailed p < .05 was considered statistically significant. First, we report the descriptive statistics on the study variables and the distributions of adults by varying severities of depression symptoms (0–5 = minimal, 6–8 = mild, 9–14 = moderate, 15–27 = severe) [25,26,38] and anxiety symptoms (0–4 = minimal, 5–9 = mild, 10–14= moderate, 15–21 = severe) [36], the maximum score is 27 for depression and 21 for anxiety. A score of PHQ-9 above 5 is considered mild depressive symptoms [35]; and mild anxiety symptoms are considered at the score of GAD-7 above 4 [36]. Second, univariate analysis (i.e. Mann-Whitney test, One-way Anova test and Kruskal-Wallis test) and an ordinal multi-class logistic regression model were used on the predictors of adults’ mental health issues [39,40].

## 3. Results

The descriptive statistics are shown in Table 1. In this sample, 45.9% of the adults were male and 54.1% were female, and the average age was 36.69 years old (SD=13.50). Over 70% (72.5%) were doing or had college degrees or above, and just under half (46.5%) were employees in their employment status. Almost half (46.7%) worked at home. The average number of children under 18 years old was 0.54. In terms of personal health status, 24.3% had some chronic health issues. The average daily exercise time in the past two weeks was 1.08 hours (SD=1.50). Additionally, 3.3% reported having the symptoms of COVID-19 infection, and 1.37 hours (SD = 1.33) on browsing information on COVID-19 online per day in average across the whole sample. Over 70% (70.3%) of the adults (N=339) had depressive symptoms (PHQ-9 score >5) and 22.8% (N=110) had experienced severe depression (PHQ-9 score = 15–27); 67.2% (N=320) had anxiety symptoms (GAD-7 score >4) and 17.2% (N=82) had experienced severe anxiety (GAD-7 score = 15–21).

Table 2 shows the univariate analysis of the screened variables. The Mann-Whitney test, One-way Anova test and Kruskal-Wallis test are used when independent variables are binary (gender and chronic health issue), continuous (age, number of children under the age of 18, exercise and hours per day browsing COVID-19 related information online) and categorical (education, employment status and experiencing symptoms of COVID-19 infection) respectively. Gender (p=0.001), age (p<0.001), education level (p=0.001), occupation (p=0.003), number of children (p=0.019), exercise (p<0.001), experiencing symptoms of COVID-19 infection (p<0.001) and hours per day browsing COVID-19 information online (p=0.009) have significant effects on depression.

**Table 2.**
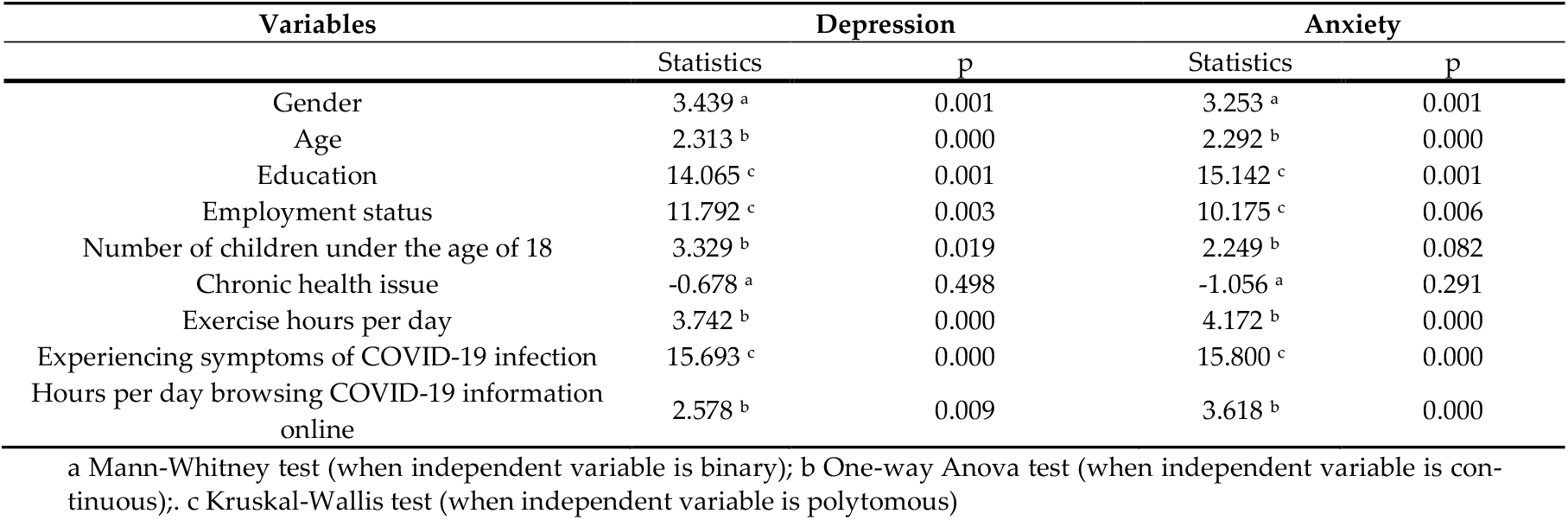
Univariate analysis of depression and anxiety.

| Variables | Depression |  | Anxiety |  |
| --- | --- | --- | --- | --- |
|  | Statistics | p | Statistics | p |
| Gender | 3.439<sup>a</sup> | 0.001 | 3.253<sup>a</sup> | 0.001 |
| Age | 2.313<sup>b</sup> | 0.000 | 2.292<sup>b</sup> | 0.000 |
| Education | 14.065<sup>c</sup> | 0.001 | 15.142<sup>c</sup> | 0.001 |
| Employment status | 11.792<sup>c</sup> | 0.003 | 10.175<sup>c</sup> | 0.006 |
| Number of children under the age of 18 | 3.329<sup>b</sup> | 0.019 | 2.249<sup>b</sup> | 0.082 |
| Chronic health issue | -0.678<sup>a</sup> | 0.498 | -1.056<sup>a</sup> | 0.291 |
| Exercise hours per day | 3.742<sup>b</sup> | 0.000 | 4.172<sup>b</sup> | 0.000 |
| Experiencing symptoms of COVID-19 infection | 15.693<sup>c</sup> | 0.000 | 15.800<sup>c</sup> | 0.000 |
| Hours per day browsing COVID-19 information online | 2.578<sup>b</sup> | 0.009 | 3.618<sup>b</sup> | 0.000 |

a Mann-Whitney test (when independent variable is binary); b One-way Anova test (when independent variable is continuous); c Kruskal-Wallis test (when independent variable is polytomous)

For anxiety, gender (p=0.001), age (p<0.001), education level (p=0.001), employment status (p=0.006), exercise (p<0.001), experiencing symptoms of COVID-19 infection (p<0.001) and hours per day browsing COVID-19 information online (p<0.001) are also significant. Contrarily, number of children and having chronic disease are non-significant (p>0.05). Considering the number of children is known from previous studies [41-43] to have a significant impact on adults’ mental health, we keep the number of children under the age of 18 in the ordinal regression model.

In an ordinal multi-class logistic regression model, the results of a parallel line test (χ2=28.835, p=0.150>0.05) showed the regression equations were parallel to each other and could be analyzed by an ordinal logistic model. The model fit was good with statistical significance (p<0.001). Similarly, the parallel line test results (χ2=21.764, p=0.474) and the degree of model fit (p<0.001) with anxiety as the independent variable were good. The results of the ordinal multi-class logistic regression are in Table 3.

**Table 3.**
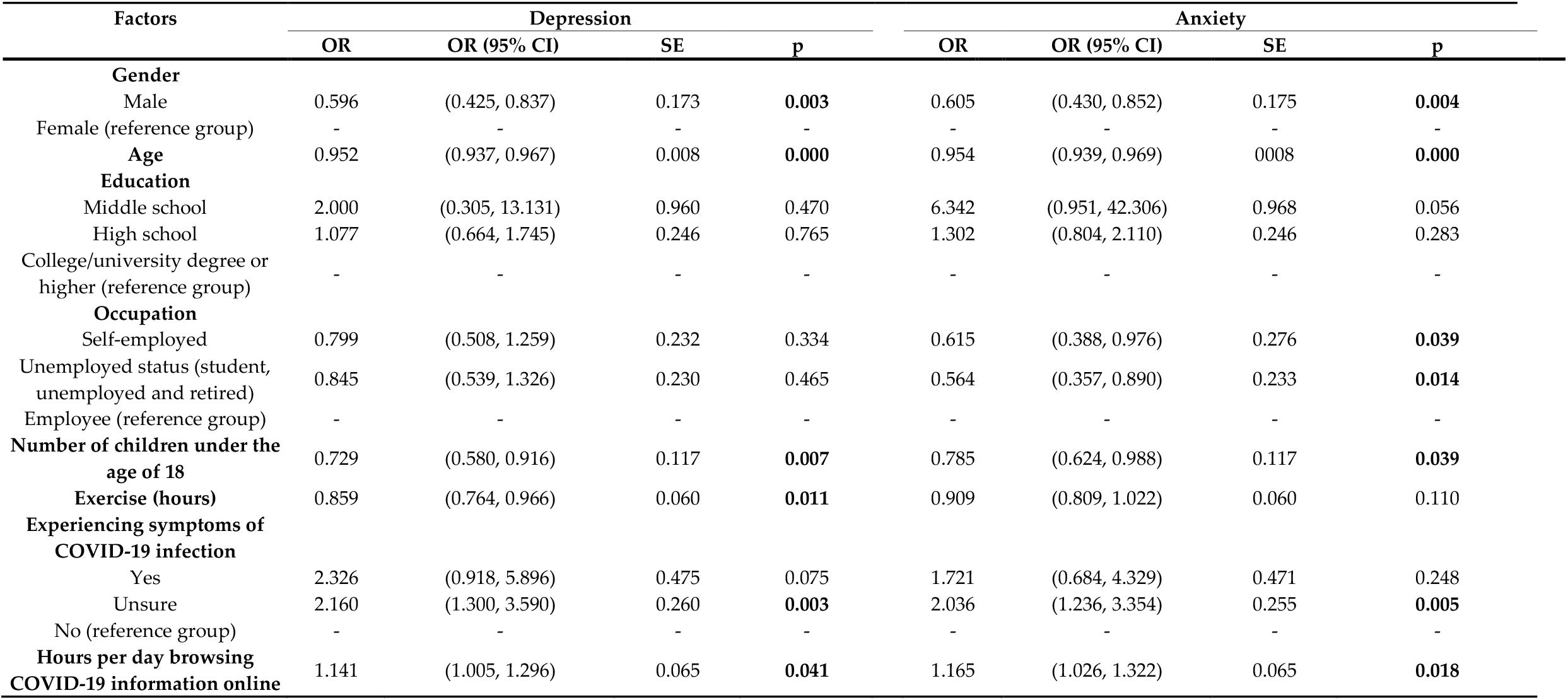
Results of ordinal multi-class logistic regression.

Table 3 reveals that males had a lower likelihood of depression during the epidemic (OR = 0.596, 95% CI = .425 – .837) than females did. Adults’ age (OR = 0.952, 95% CI = .937 – .967), number of children (OR = 0.729, 95% CI = .580 – .916) and daily exercise time (OR = 0.859, 95% CI = .764 – .966) negatively predicted depression, and adults who were unsure whether they had experienced symptoms of COVID-19 infection were more likely to experience depression (OR = 2.160, 95% CI = 1.300 – 3.590). Hours per day browsing COVID-19 information online predicted depression positively (OR = 1.141, 95% CI = 1.005 – 1.296).

Similarly, males were less likely to experience anxiety than females were (OR = 0.619, 95% CI = .438 – .876). Age (OR = 0.954, 95% CI = .939 – .969) and the number of children (OR = 0.785, 95% CI = .624 – .988) negatively predicted anxiety. Additionally, people who were unsure whether they had COVID-19 infection (OR = 2.036, 95% CI = 1.236 – 3.354) and who spent more time browsing COVID-19 information online (OR = 1.165, 95% CI = 1.026 – 1.322) were more likely to have anxiety symptoms. Other variables in the model, such as education, had no significant predictive effect on either depression or anxiety.

## 4. Discussion

The COVID-19 pandemic has had a massive impact on people’s lives, especially in Brazil due to the limited health system capacity to deal with the COVID-19 crisis [44]. Nonetheless, to date, few studies have examined the mental health conditions of adults in Brazil, which leads the world in daily COVID-19 cases and death in 2020. In our study, close to half of the adults were unable to work in their normal workplaces. The results of our survey of adults in Brazil reveal the prevalence of depressive symptoms was 70.3%, and the incidence of severe depressive symptoms was 22.8%; the incidence rates of anxiety symptoms and severe anxiety symptoms were 67.2% and 17.2% respectively. Several recent studies before COVID-19 reported the incidence of anxiety in Brazilians was 18.0% in year 2018 [45] and the average incidence of depression in Brazilians was 4.1% in year 2013 [46], which were much lower than our results which are the prevalence rates after the initial COVID-19 outbreak. Given Brazil is the largest country in South America, to better benchmark and interpret our findings, we list the major mental health studies in Latin America during the COVID-19 pandemic to date to provide more comprehensive evidence on the mental burden among Brazilian adults (see Table 4). The table reveals the prevalence rates in our study in Brazil are higher than many studies in other South American countries such as 47.0% prevalence of depressive symptoms and 54.9% of anxiety symptoms in Argentina [47], 19.2% prevalence of psychological distress in Chile [48], 34.9% prevalence of depressive symptoms [49], and 21.7% of severe anxiety symptoms and 26.1% of severe mental distress in Peru [2].

**Table 4.** Prevalence rates of mental issues during the COVID-19 pandemic in South America.

| Study | Duration | Country | Sample | Instruments and cut-off point | Mental health indicators |
| --- | --- | --- | --- | --- | --- |
| Torrente et al. (2020) [47] | March 24 to March 27, 2020 | Argentina | Adults (N=10,053) | PHQ-9 (6, 9, and 15 as the cut-off points)<br>GAD-7 (5, 10, and 15 as the cut-off points) | 47.1% prevalence of **anxiety** symptoms (18.5% mild, 18.1% moderate and 10.5% severe symptoms)<br>54.9% prevalence of **depressive** symptoms (31.6% mild, 13.6% moderate and 9.6% severe symptoms) |
| Duarte & Jiménez-Molina (2020) [48] | Between May and June 2020 | Chile | Adults (N=1078) | PHQ-4 (6 as the cut-off point of prevalence) | 19.2% prevalence of **psychological distress** |
| Herrera et al. (2020) [50] | Between June and September 2020 | Chile | Older adults (N=720) | PHQ-9 (6, 9, and 15 as the cut-off points)<br>Geriatric Anxiety Inventory – Short Form (GAISF) (3 as the cut-off point of prevalence) | 30.18% prevalence of **depressive** symptoms<br>42.85% prevalence of **anxiety** symptoms |
| Paz et al. (2020) [51] | March 22 to April 18, 2020 | Ecuador | Confirmed or suspected COVID-19 patients (N=759) | PHQ-9 (6, 9, and 15 as the cut-off points)<br>GAD-7 (5, 10, and 15 as the cut-off points) | 20.3% prevalence of **depressive** symptoms<br>22.5% prevalence of **anxiety** symptoms |
| Chen et al. (2020) [52] | April 10 to May 2, 2020 | Ecuador | Healthcare workers (N=252) | GAD-7 (5, 10, and 15 as the cut-off points)<br>K-6 (5, 13 as the cut-off points) | 32.5% prevalence of **distress disorder**<br>28.2% prevalence of **anxiety** symptoms |
| Romero Parra (2020) [53] | Do not report | Peru, Venezuela | University students (N=600) | Beck Depression Inventory (BDI-2) (14, 20, and 29 as the cut-off points) | 34.7% prevalence of **depressive** symptoms of university students in Peru (18.4% mild, 11.2% moderate, 5.1% severe symptoms)<br>11.4% prevalence of **depressive** symptoms of university students in Venezuela (6.4% mild, 5.0% moderate symptoms) |
| Antiporta et al. (2021) [49] | May 4 to May 16, 2020 | Peru | Adult Peruvian residents (N=57,446) | PHQ-9 (6, 9, and 15 as the cut-off points) | 34.9% prevalence of **depressive** symptoms |
| Yañez et al. (2020) [2] | April 10 to May 2, 2020 | Peru | Healthcare workers (N=303) | GAD-7 (5, 10, and 15 as the cut-off points)<br>K-6 (5, 13 as the cut-off points) | Mean of GAD-7 **anxiety** scale is 15.4<br>Mean of K6 **distress** scale is 19.2<br>21.7% prevalence of **severe anxiety** symptoms<br>22.5% prevalence of **severe mental distress** |
| Martínez et al. (2020) [54] | April 2020 | Colombia | Adults, college students, and informal workers (N=984) | Not reported | Mean of **anxiety and stress** score is 6.5 (scale 0–10)<br>Mean of **depressed** score is 3.8 (scale 0–10) |
| Zhang et al. (2021) [21] | March 25 to March 28, 2020 | Brazil | Adult Brazilian residents (N=638) | COVID-19 Peritraumatic Distress Index (CPDI) (4, 28, and 52 as the cut-off points) | Mean score of COVID-19 Peritraumatic Distress Index (CPDI) was 37.64 (score 0–100)<br>71.8% prevalence of **peritraumatic distress** (52.0% mild or moderate, 18.8% severe distress) |

Our regression analysis reveals that females had higher likelihood of depression and anxiety symptoms, a finding that is in line with the previous research on gender and mental health [55]. Older adults and those with more children were less likely to experience anxiety and depression symptoms [56]. Adults who exercised more per day during the pandemic had a lower likelihood of depression symptoms, supporting the view that exercise might help relieve the buildup of depressive symptoms [57]. Employees, compared with the unemployed or self-employed, were more likely to have anxiety but not depressive symptoms. Such a finding differs from the literature that suggests occupational stressors may cause both depression and anxiety in existing studies [58]. Our findings on the higher anxiety symptoms of employees suggest COVID-19 may present some unique challenges on anxiety across employment status groups. The higher anxiety symptoms experienced by employees might be related to the drastic changes in the work environment during COVID-19, while self-employed individuals and non-working groups do not have such trouble. In addition, a potential reason why our study did not find an association between occupation and anxiety is that not all kinds and levels of occupational stress might carry a significant relationship with depressive symptoms [59].

Individuals’ hours per day browsing COVID-19 information online positively predicted depression and anxiety, suggesting online information on crises might exacerbate mental disorders [60-61]. Altogether, our findings identified several predictors which enable psychiatrists and healthcare organizations to better identify and focus on the more vulnerable sub-populations. Furthermore, our results may enable psychiatry practitioners to identify potential patients with depressive and anxiety symptoms during the COVID-19 pandemic.

## 5. Limitation and future research

The study has certain limitations. Firstly, we designed a cross-sectional study, which yields a snapshot rather than a dynamic picture, and we suggest longitudinal designs in future research. Another potential limitation regarding our sampling procedures is that although the sample covered much of Brazil geographically, it was not entirely representative of the population due to our online survey, because only 71% of the population has access to the Internet in Brazil. Our small sample size may raise concerns about generalizability. In future research it would be especially interesting to investigate populations that do not have internet access. Although it was proposed that web surveys had an 11 percentage points lower response rate than other survey modes [62], we still cannot ignore the non-response bias problem that may be caused by low response rates in this study (56.2%). Future research can focus on increasing the response rate and sample size to extend our findings. Brazil is a very large country, and research on mental health during COVID-19 in Latin America remains underdeveloped, calling for more research to generate evidence to better cope with the ongoing pandemic [63]. We hope our results help to gather data for evidence-based decisions on mental health for Brazil, one of the largest and worst-affected countries in the COVID-19 crisis.

## 5. Conclusions

This study reported 70.3% prevalence of depressive symptoms (17.2% mild, 30.3% moderate and 22.8% severe symptoms) and 67.2% prevalence of anxiety symptoms (31.3% mild, 18.7% moderate and 17.2% severe symptoms) among Brazilian adults (N=482) during the COVID-19 pandemic. Gender, age, number of children, being employed and time browsing COVID-19 information online are potential predictors of experiencing depressive and anxiety symptoms. The incidence of anxiety and depressive symptoms in the Brazilian adult population was much higher after the initial outbreak than the pre-pandemic rates, indicating Brazilians’ mental health has suffered during the COVID-19 pandemic. Healthcare organizations can use our findings to identify groups mentally vulnerable to COVID-19 in Brazil. Better identification of the mentally vulnerable population can enable more targeted effort to reduce the high prevalence of mental health symptoms in Brazil. Beside psychiatric identification and resource prioritization, policy-makers can direct and promote more reliable information on the pandemic online, which has been a source of mental health issues in the pandemic. The findings of this study quantify the prevalence rates of depression and anxiety symptoms in Brazil and identify several predictors, which can enable psychiatrists and healthcare organizations to better identify the more vulnerable sub-populations and provide evidence to deploy resources as well as create opportunities for timely pre-emption and prevention.

## Data Availability

Data are available upon request.

## Author Contributions

Conceptualization, S.X.Z. and H.H.; methodology, S.X.Z. and H.H.; software, S.X.Z. and H.H.; validation, S.X.Z. and H.H.; formal analysis, S.X.Z. and H.H.; investigation, S.X.Z., H.H. and J.L.; resources, S.X.Z., H.H. and J.L.; writing—original draft preparation, S.X.Z. and H.H.; writing—review and editing, S.X.Z. and H.H.; supervision, S.X.Z.; visualization, H.H.; data curation, M.A., S.F.P. and J.A.S.; project administration, S.X.Z. and J.A.S. All authors have read and agreed to the published version of the manuscript. Authorship must be limited to those who have contributed substantially to the work reported.

## Informed Consent Statement

Informed consent was obtained from all subjects involved in the study.

## Data Availability Statement

Data are available upon request.

